# Proteo-genomic analysis of SARS-CoV-2: A clinical landscape of SNPs, COVID-19 proteome and host responses

**DOI:** 10.1101/2020.11.27.20237032

**Authors:** Sheetal Tushir, Sathisha Kamanna, Sujith S Nath, Aishwarya Bhat, Steffimol Rose, Advait R Aithal, Utpal Tatu

## Abstract

A novel severe acute respiratory syndrome coronavirus 2 (SARS-CoV-2) is the causative agent of COVID-19 and continues to be a global health challenge. To understand viral disease biology, we have carried out proteo-genomic analysis using next generation sequencing (NGS) and mass-spectrometry on nasopharyngeal swabs of COVID-19 patients to examine clinical genome and proteome. Our study confirms the hyper mutability of SARS-CoV-2 showing multiple SNPs. NGS analysis detected 27 mutations of which 14 are synonymous, 11 are missense and 2 are extragenic in nature. Phylogenetic analysis of SARS-CoV-2 isolates indicated their close relation to Bangladesh isolate and multiple origins of isolates within a country. Our proteomic analysis, for the first time identified 13 different SARS-CoV-2 proteins from the clinical swabs. Of the total 41 peptides captured by HRMS, 8 matched to nucleocapsid protein, 2 to ORF9b, 1 to spike glycoprotein and ORF3a, with remaining mapping to ORF1ab polyprotein. Additionally, host proteome analysis revealed several key host proteins to be uniquely expressed in COVID-19 patients. Pathway analysis of these proteins points towards modulation in immune response, especially involving neutrophil and IL-12 mediated signaling. Besides revealing the aspects of host-virus pathogenesis, our study opens new avenues to develop better diagnostic markers and therapeutics.

## INTRODUCTION

Coronavirus disease 2019 (COVID-19) is the latest addition to the extensive list of infectious diseases caused by viruses that have jumped from animals to humans. The global outbreak of COVID-19 caused by severe acute respiratory syndrome coronavirus 2 (SARS-CoV-2) originated from the city of Wuhan, China in December 2019. After SARS-CoV in 2003 ^1,2^ and Middle East respiratory syndrome coronavirus (MERS-CoV) in 2012, ^3^ SARS-CoV-2 is the third of coronavirus family to cross the species barrier and infect humans with severe respiratory disease. It has proven to be more dangerous than SARS-CoV and MERS-CoV due to its alarming transmission rate through respiratory droplets, encountering infected persons or contaminated surfaces.^4^ SARS-CoV-2 was first identified by meta-transcriptomic sequencing from the bronchoalveolar lavage fluid of a patient in China and the sequence was made available at Global Initiative on Sharing All Influenza Data (GISAID) platform on 12^th^ January 2020.^5^ Phylogenetic analysis shows that about 30 kb genome of this new RNA virus is most closely related to a group of SARS-like coronavirus (humans and bat) with 89.1% similarity.^5^ SARS-CoV-2 has 14 open reading frames (ORF), which codes for structural, accessory non-structural and replication proteins. ORF1ab is the largest of all having 21,291 nucleotides and codes for replicase polyprotein. Structure protein genes located downstream to ORF1ab, aligned in the following order-spike (S), envelope (E), membrane (M) and nucleocapsid (N) with ORFs that code for accessory non-structural proteins are located in between. SARS-CoV-2 forms a capsid of lipid bilayer with embedded E, M and S proteins.^6^ Till the time any drug or efficient vaccines are developed, the biggest challenge in front of the scientific committee and health workers is to contain the spread of the virus by testing, tracing and isolation of infected subjects until recovery.

Since the first SARS-CoV-2 sequence was made available, more than 140,000 complete genome sequences have been added to the list by laboratories across the world. Despite a large number of sequences being available, it is still not clear how fast the virus mutates and if the mutations impact its virulence in the context of the growing pandemic. In addition, very little information is available regarding the clinical proteome of the virus. Until recently, only a few proteins, mainly including structural proteins N and S have been identified from clinical swabs. Besides, host proteome studies from clinical samples are necessary to fill the void in understanding the host response to viral infection.

In this study, genomic analysis of COVID-19 was performed by next generation sequencing (NGS) on swab samples of Reverse transcription-polymerase chain reaction (RT-PCR) positive individuals, collected from Bangalore, India. Variant analysis of these samples showed a high mutation rate, with ≥11 mutations observed per sample. Phylogenetic analysis of these sequences with other variants from India as well as across the world revealed their close relation to one of the Bangladesh isolates, which had showed European origin. SARS-CoV-2 phylogeny indicated the prevalence of isolates showing multiple origins within a country. Overall, through genomic analysis of SARS-CoV-2, our study highlighted increasing variations (SNPs) in the viral genome and their role to understand its evolution and virulence.

In addition to sequencing the genome by NGS, our study also explored the COVID-19 clinical proteome and host-protein responses by using high-resolution mass spectrometry (HRMS). We performed HRMS on swab samples of both RT-PCR positive and negative patients. In total, we identified 41 peptides matching to 13 different COVID-19 proteins, including proteins from ORF1ab polyprotein, Spike glycoprotein, ORF3a, ORF9b and Nucleocapsid. Additionally, the host proteomic analysis revealed significant differences between RT-PCR positive and RT-PCR negative host proteomes. We found 441 proteins to be uniquely present in positive samples. Most of these proteins are involved in neutrophil degranulation and activation pathways indicating host immunological response to the virus. In conclusion, our analysis confirms the presence of COVID-19 peptides in nasal swab samples and proteomic analysis also predicted host responses to viral infection, including identification of the neutrophil response as a key host response against COVID-19 infection.

## MATERIALS AND METHODS

### Sample Collection

Nasopharyngeal swab samples were collected from the diagnosed patients as a part of routine monitoring. Part of the samples after diagnosis were sent to the lab for research purpose. Samples were classified into positive and negative based on the RT-PCR result targeting E and RNA-dependent RNA polymerase (RdRp) genes of the virus. Samples were collected only after the approved consent of the patients who were informed about the study. The study was conducted after approval of the institutional human ethics committee, IISc (19-01092020).

### RNA Library Preparation

Total RNA from nasopharyngeal swabs of three positive patients (RT-PCR test) was extracted using Trizol based extraction. RNA samples were quantified using Qubit RNA Assay HS (Invitrogen). RNA purity was checked using Nanodrop and integrity was assessed on TapeStation using RNA HS ScreenTapes (Agilent). Qiagen SARS-CoV-2 Primer (Qiagen) was used to prepare libraries from RNA extracted from COVID-19 positive subjects. Viral RNA was converted to cDNA and used as a template for multiplex PCR with primers spanning the entire genome of the virus. The amplicons were then pooled and purified before proceeding for library preparation. During library preparations, the amplicons were subjected to a series of enzymatic steps that repair the ends, tails the 3’ end with a single ‘A’ nucleotide, followed by ligation of the adapters. The adapter-ligated products were then purified and enriched using a limited cycle PCR. The final cDNA libraries were purified and checked for fragment size distribution on TapeStation using D1000 DNA ScreenTapes (Agilent).

### Next-generation sequencing of SARS-CoV-2

Prepared cDNA libraries were quantified using Qubit High Sensitivity Assay (Invitrogen). Quantified libraries were pooled and diluted to final optimal loading concentrations for cluster amplification on Illumina flow cell followed by sequencing on Illumina HiSeq X instrument to generate 150 bp paired-end reads.

### Mutation analysis

Initially, the quality of the reads was checked using FastQC v0.11.9.^7^ Further, the sequencing adapters clipped at 5’ and 3’ end of reads were trimmed using Cutadapt v2.9.^8^ The adapter trimmed pair-end reads were then aligned to the Wuhan reference genome (Accession No. NC_045512.2) downloaded from NCBI.^9^ The fast and accurate read alignment was achieved by using BWA v.0.7.12 aligner.^10^ The aligned reads were sorted, removed soft-clippings and then variant calling was performed using GATK variant caller.^11^ The reported variants were then annotated to study their effects in proteins and genes using SNPEff tool.^12^ The variant class, amino acid changes and other relevant annotations were added to the variants.

### Nucleotide Sequence Accession Number

The SARS-CoV-2 whole genome sequences have been submitted to NCBI under the accession number PRJNA668889.

### Phylogenetic Analysis

The evolutionary history was inferred by using the Maximum Likelihood method and the Tamura-Nei model.^13^ The tree with the highest log likelihood (−525441.09) was generated. The initial tree for the heuristic search was obtained automatically by applying Neighbor-Join and BioNJ algorithms to a matrix of pairwise distances estimated using the Tamura-Nei model and then selecting the topology with superior log likelihood value. The tree was drawn to scale, with branch lengths measured in the number of substitutions per site. The analysis involved 40 nucleotide sequences with a total of 29945 positions in the final dataset. Codon positions included were 1st+2nd+3rd+Noncoding. Evolutionary analyses were conducted in MEGA X.^14^

### Sample preparation for Mass spectrometry

In-solution sample preparation was done to extract the peptides from the protein solution. Briefly, samples collected in VTM were centrifuged at 14,000 rpm for 15 min at 4°C. The supernatant was collected in a separate microcentrifuge tube and the pellet was washed with 1x PBS. Further, the pellet containing epithelial cells was lysed by 1x Triton buffer (chilled). Lysate in the supernatant was collected after centrifuging at 14,000 rpm for 20 min at 4°C. Both lysate and supernatant were proceeded for protein precipitation by the addition of chilled acetone and kept at −80°C for 2 hrs. Precipitated proteins were washed with chilled acetone and dissolved in 50mM ammonium bicarbonate. Proteins were reduced using 10mM DTT (Sigma-Aldrich) in 50mM ammonium bicarbonate at 56°C for 45 mins followed by alkylation with 55mM iodoacetamide in 50mM ammonium bicarbonate at 37°C for 30 mins in the dark. In-solution digestion was carried out by adding trypsin (Promega) 1µg/µl to a final protease to protein ratio of 1:50 (w/w) and incubated at 37°C for 16 hours, with frequent shaking. Digestion was stopped using formic acid and all the samples were subjected to vacuum dry.

### Mass Spectrometry and Database Search

The dried trypsin digested peptides were reconstituted in a mixture of 20% ACN and 80% MQ containing 0.01% formic acid. The protein digests were analyzed using Agilent 1290 Infinity II LC system coupled with Agilent Advance Bio Q-TOF (6545XT). The column used for chromatography was Agilent AdvanceBio Peptide Map (2.1x 150mm, 2.7µ). Mobile phase A was MQ (0.1% formic acid) and mobile phase B was ACN (0.1% formic acid). The peptides were separated by using a 90 min gradient flow at a flow rate of 0.4 ml/min. The MS and MS/MS scan were acquired in the positive mode and stored in centroid mode. The following MS data acquisition parameters, Vcap was set at 3500V, drying gas flow rate and the temperature was set at 12 L/min and 270°C, respectively. Collision energy with a slope of 3.6V/100 Da and an offset of 4.8V was used for fragmentation. The precursor ion data were captured in a mass range of 200-1800 m/z and product ions data were acquired in the range 50-2900 m/z. Reference exclusion was given for 0.05min after 1 spectrum. The raw data were analyzed using MaxQuant software (v1.6.2.10) and processed through the MS excel sheet. The database analysis was performed against SARS-CoV-2 proteome and Homo sapiens proteome in the UniProt database (Proteome ID-UP000005640). The following search parameters were used for the database analysis: Precursor mass tolerance: 10ppm, fragment mass tolerance: 40ppm with cysteine carbamidomethylation as a fixed modification and methionine oxidation and protein N-term acetylation as variable modifications.

### MS Data repository

Mass spectrometry proteomics data acquired on nasopharyngeal swabs have been deposited to the ProteomeXchange Consortium via the PRIDE^15^ partner repository under dataset identifiers PXD021896 and 10.6019/PXD021896.

### Gene ontology and Pathway Analysis

All Uniprot IDs of host proteins found exclusively in COVID-19 positive samples were extracted and analyzed through DAVID Tool for conversion to Entrez IDs. These Entrez IDs were then used for the identification of Gene Ontology terms and pathways. Statistical significance of the genes involved in positive samples was analyzed using the R package, clusterProfiler. To determine whether any terms annotate a specified list of genes at a frequency greater than that would be expected by chance, clusterProfiler calculates a p-value using the hypergeometric distribution. Statistically enriched GO terms were then plotted and analyzed through dot plot, category net plot (Cnetplot) and enrichment map (Emap).

## RESULTS

### Genome sequence reveals emerging mutations in SARS-CoV-2

Since the first SARS-CoV-2 genome sequence was shared on 12^th^ Jan 2020, ^5^ there are more than 133,000 genome sequences are available at GISAID to date.^16^ To correlate the sequence of prevailing COVID-19 with those reported earlier, we carried out Illumina HiSeq X, NGS of SARS-CoV-2. COVID-19 RNA from nasopharyngeal swabs, tested positive by RT-PCR was converted to cDNA and processed for NGS as described in the methods section. NGS analysis retrieved the complete genome sequence from all three samples. FastQ files generated from NGS were reference mapped to SARs-CoV-2 isolate, Wuhan-Hu-1 (Accession No. NC 045512.2) with 100% genome coverage. The alignment of these 3 isolates with reference genome showed the prevalence of single nucleotide polymorphism (SNPs) (Figure1). Sample 1, 2 and 3 showed 11, 16 and 19 SNPs, respectively. The number of mutations observed are higher than the average mutations per sample of around 7 worldwide.^17^ A total of 27 variations were found in isolates out of which 4 are common to all and 11 are exclusively common to samples 2 and 3. The 4 common mutations observed in all three isolates are c.241C>T, c.3037C>T, c.14408 C>T and c.23403A>G. In total, we found 9 mutations belonging to the category of most frequent mutations out of which 6 (c.241C>T, c.3037C>T, c.14408C>T, c.23403A>G, c.25563G>T and c.28881G>A) are common in all the continents, c.26735C>T and c.28854C>T are specific to Asian isolates and c.18877C>T found in both America and Asian isolates.

The amino acid substitutions due to these point mutations are represented in the lower panel of Figure 1. Out of 27, 25 mutations are in the coding region, which generate 14 synonymous and 11 missense amino acid substitution. 3 of these 11 missense mutations could be of high impact as they substitute charged to uncharged amino acid or vice-versa, hence, may impact the structure and consequently the function of the proteins. These include-p.D614G in Spike glycoprotein, p.Q57H in ORF3a and p.G204R in Nucleocapsid.

**Figure 1.**
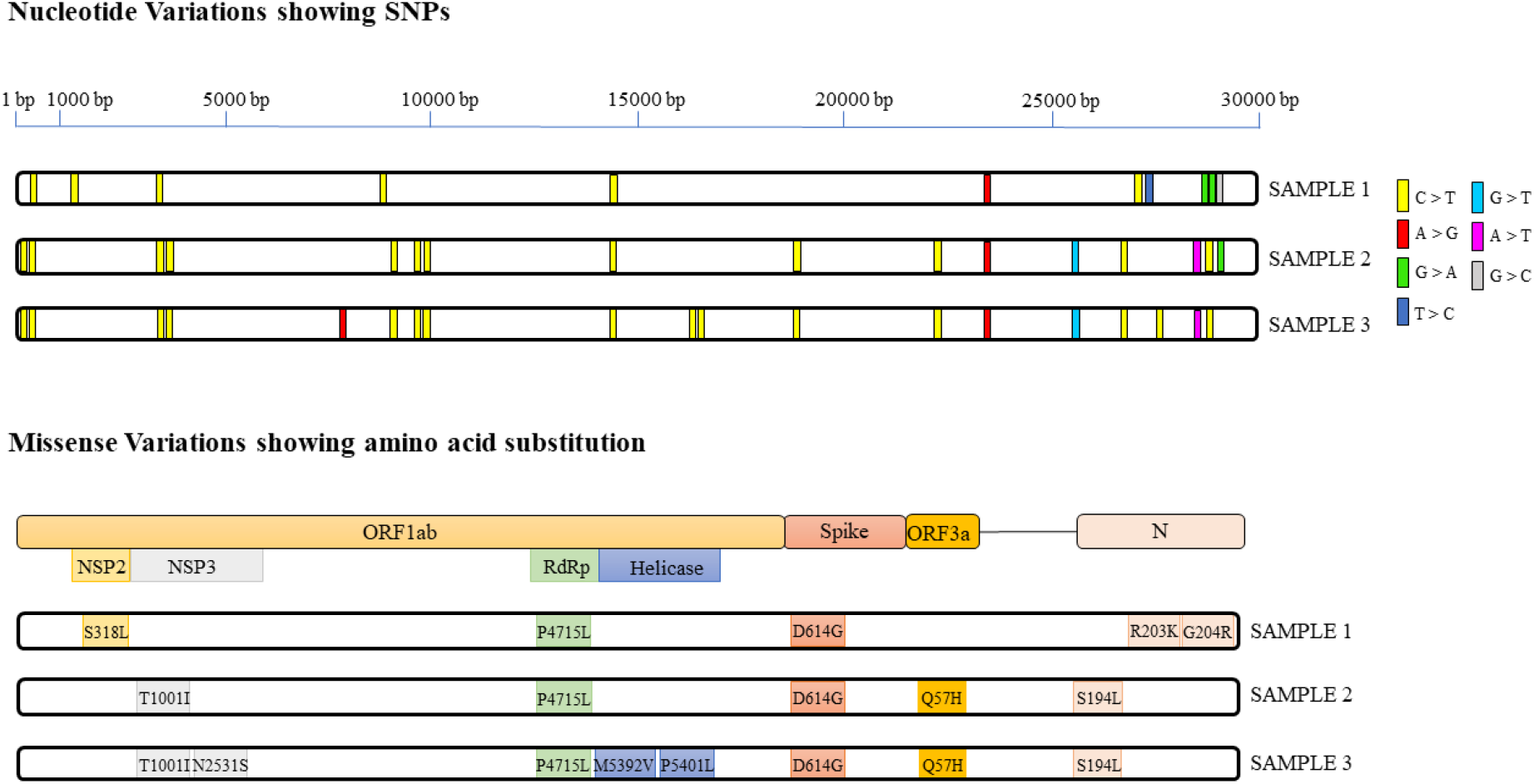
Variant analysis in Indian isolates of SARS-CoV-2 genome. The top panel of the figure indicates nucleotide variations (SNPs) in the genome sequence of the SARS-CoV-2 Bangalore isolates against the reference Wuhan-Hu-1 isolate complete genome sequence. Specific transitions and transversions with their color coding are mentioned on the right side of the panel. The bottom panel indicates position of the missense mutations against the reference Wuhan-Hu-1 SARS-CoV-2 isolate.

**Figure 2.**
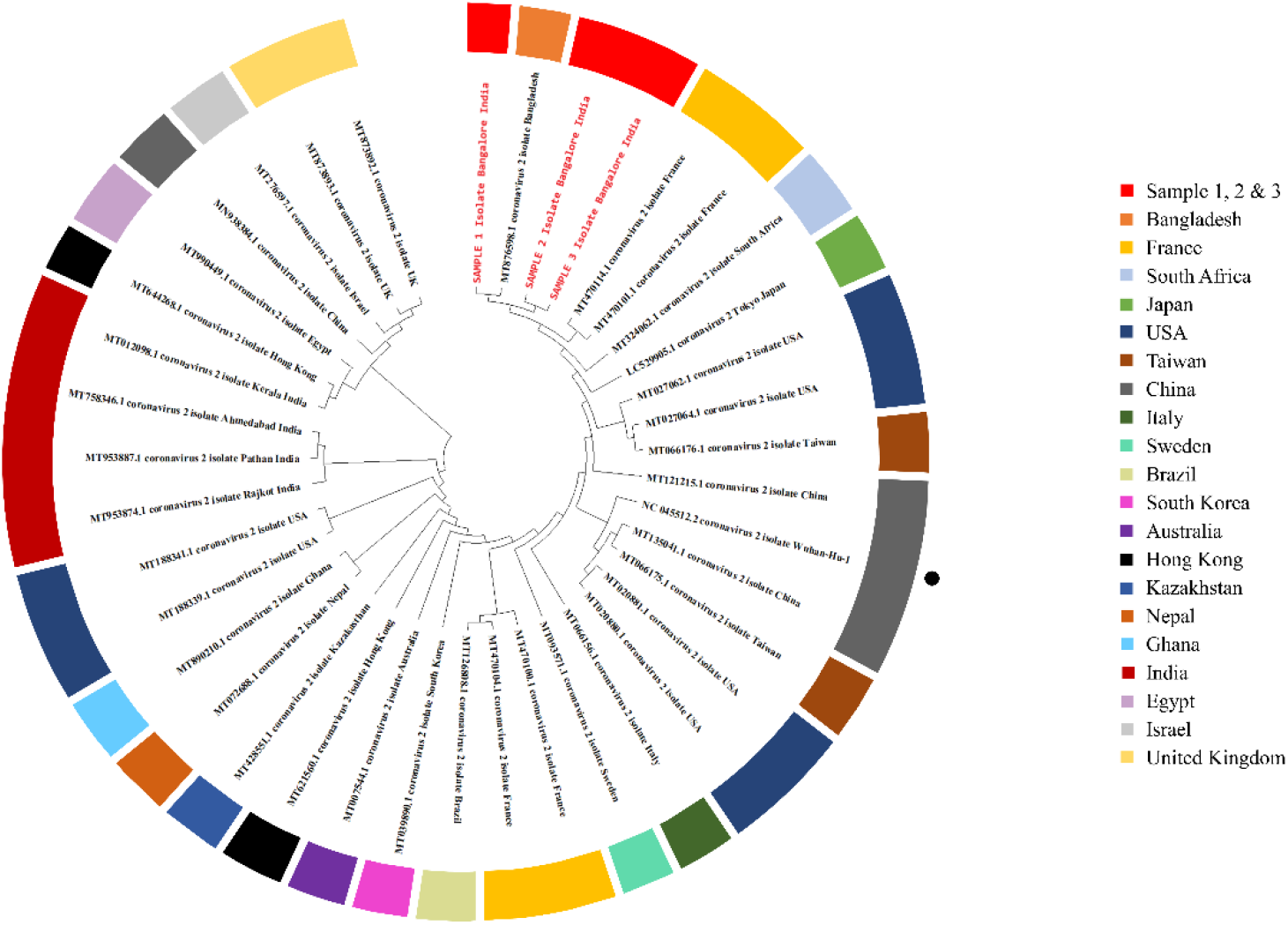
Phylogenetic analysis of SARS-CoV-2 isolates. Whole genome phylogeny representing relationship of Bangalore SARS-CoV-2 isolates based on Maximum Likelihood method and Tamura-model created using MEGA X. The phylogenetic analysis involved 40 SARS-CoV-2 sequences representing variants from 20 countries around the globe. The colors around the tree represents country of origin for each isolate. Isolates from Bangalore are represented in red text showing close relation to Bangladesh isolate. Black dot at outer region of the circle marks the Wuhan-Hu-1 reference genome.

Further, to see the evolutionary relationship of these isolates we constructed a phylogenetic tree using MEGA X software. Phylogenetic analysis showed that sample 2 and 3 are more closely related and all three are in close relation to one of the Bangladesh isolates, which appears to be originated from France isolates. Distinct clade-wise assignment revealed that all the isolates belong to G derived clade (European origin), isolate 1 to GR and isolate 2 and 3 to GH.

### Clinical proteome of SARS CoV-2

Despite an ample collection of genome information, the information on clinical proteome of COVID-19 is ill explored. Only around 15 studies reported the proteome of SARS-CoV-2.^18^ We carried out HRMS analysis of SARS-CoV-2 clinical proteome. Part of the nasopharyngeal samples obtained from 12 COVID-19 positive patients were analyzed using Agilent Advance Bio Q-TOF (6545XT). Extracted proteins from swabs were reduced, alkylated and digested as described in the methods section. From MS/MS spectra recorded at FDR ≤1% we were able to identify 41 unique peptides matching to 13 different viral proteins. The maximum number of peptides were attributed to ORF1ab polyprotein. As showed in peptide-map in Figure 3, we detected 8 peptides matching to Nucleocapsid protein, 7 peptides to NSP3, 6 peptides to exoribonuclease, 4 peptides to 2’-O-methyltransferase, 3 peptides each to RdRp (NSP12) and endoribonuclease, 2 peptides each to NSP2, Helicase and protein 9b and 1 peptide each to NSP8, NSP10, Spike glycoprotein and protein 3a. Peptide ITEHSWNADLYK (2’O Methyltransferase) was detected in 50% of the sample (6/12) and peptide IVQMLSDTLK (exoribonuclease) detected with the highest intensity. Sample 12 showed the maximum number of identified peptides (8 attributed to 6 different proteins). Apart from sample 1,3,10 and 11, peptides for 2’-O-methyltransferase protein are detected in all the sample (66.6%). Our MS result showed a correlation with the result of RT-PCR for positive samples. Our result suggests the potential of mass spectrometry for highly sensitive and reliable diagnosis.

**Figure 3.**
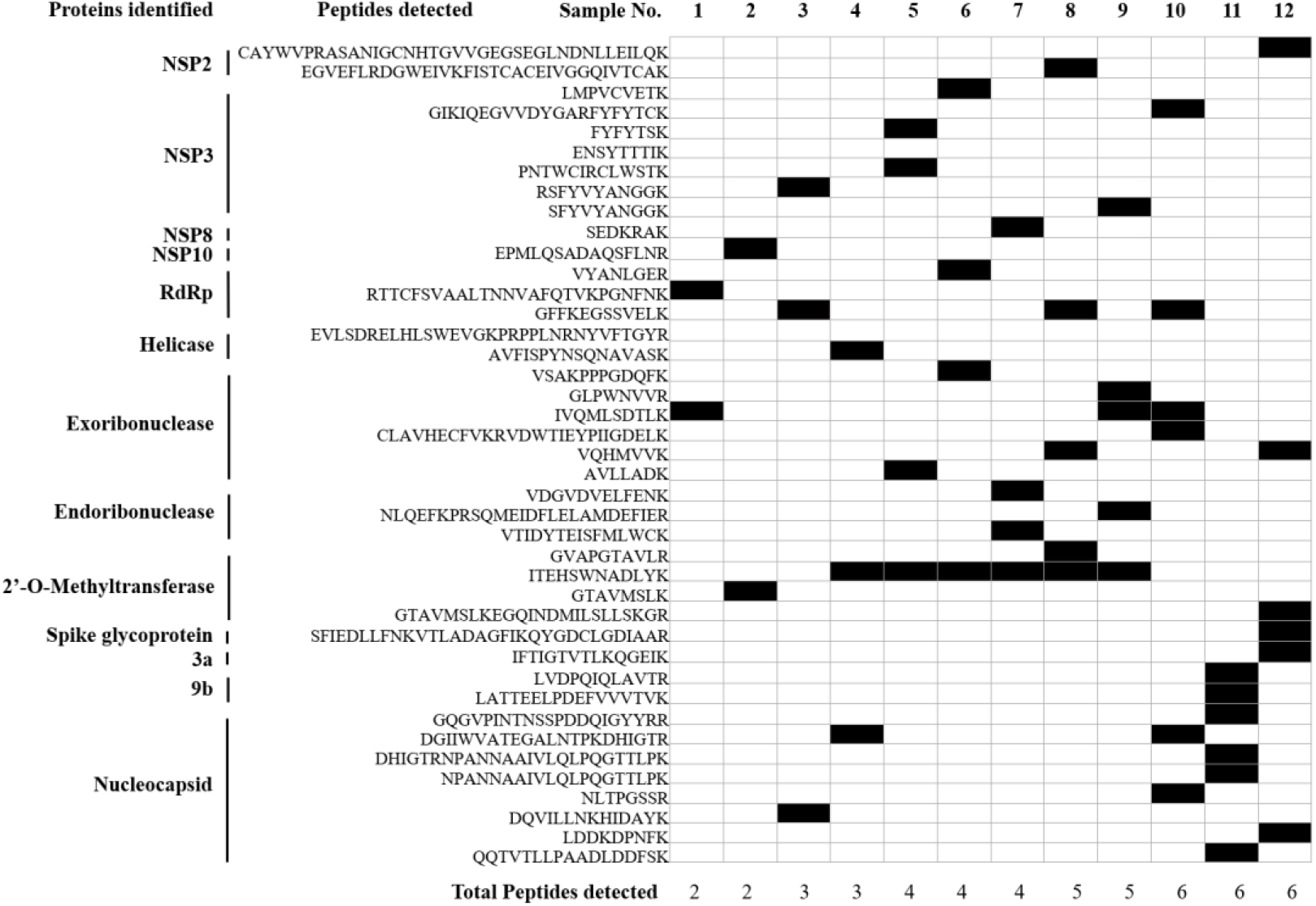
Clinical proteome of SARS-COV-2. Peptide map depicts COVID-19 peptides identified from the clinical nasopharyngeal swabs of COVID-19 patients. Cells highlighted in black represent detected peptides in that particular sample. Sequence of the peptides along with the matched protein are listed on the left. Total peptides identified in the sample are indicated at the bottom. The sample numbers 1-12 are arranged in increasing order of total peptides detected.

### Host responses to SARS CoV-2 infection

We also looked for host protein dynamics upon SARS-CoV-2 infection by searching the MS data against the human proteome database (Proteome ID-UP000005640). For this, we analyzed 9 samples of both COVID-19 positive and negative patients. Negative being those which tested negative by RT-PCR and which did not show any COVID-19 peptides. To characterize the pathways that are getting modulated by the viral infection, we compared the list of host proteins identified by LC-MS/MS in all positive and negative samples. Figure 4A represents the Venn diagram of host proteins. We identified 441 proteins to be uniquely present in positive sample, 246 exclusively in negative sample and 158 found common to both.

**Figure 4.**
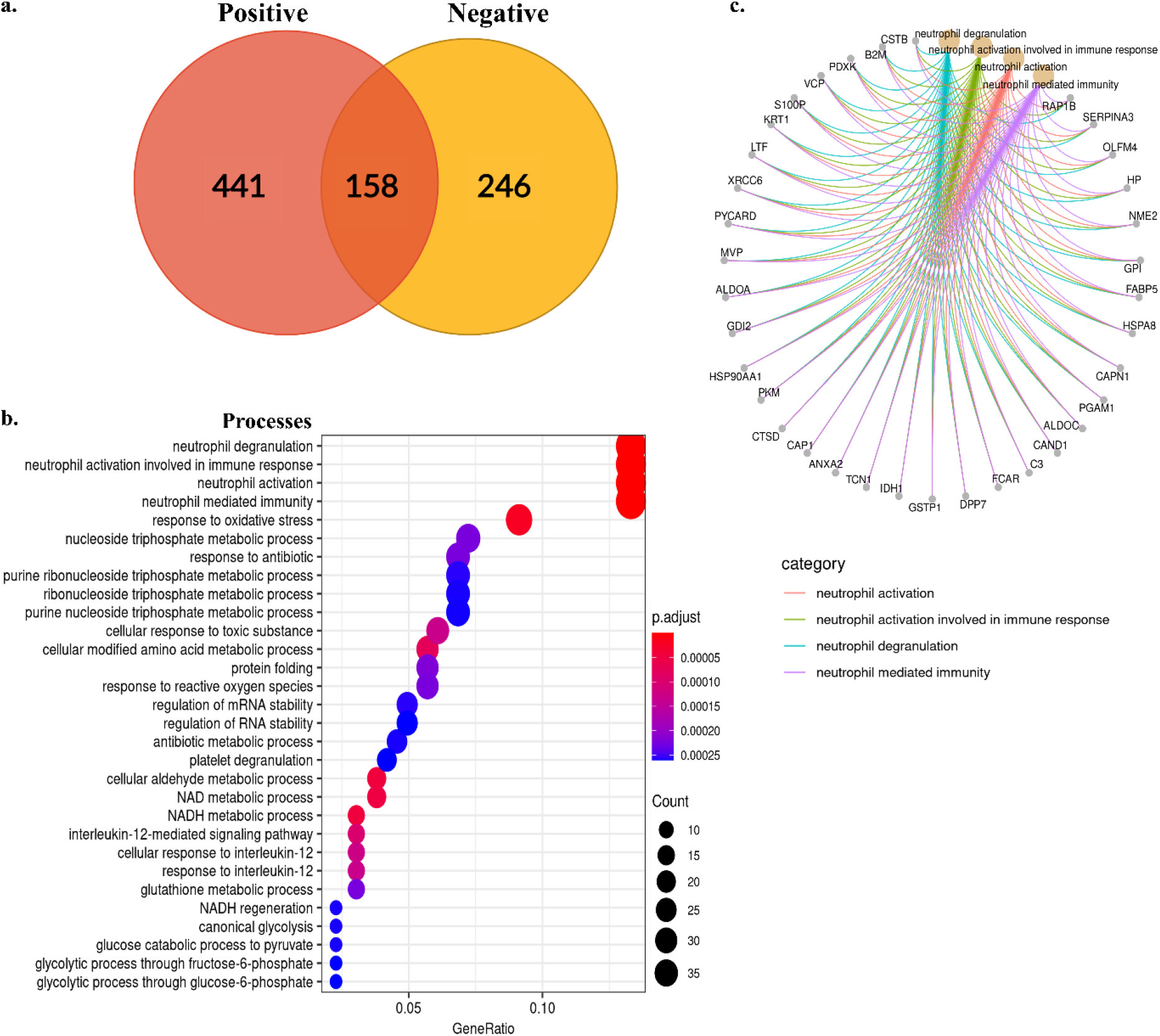
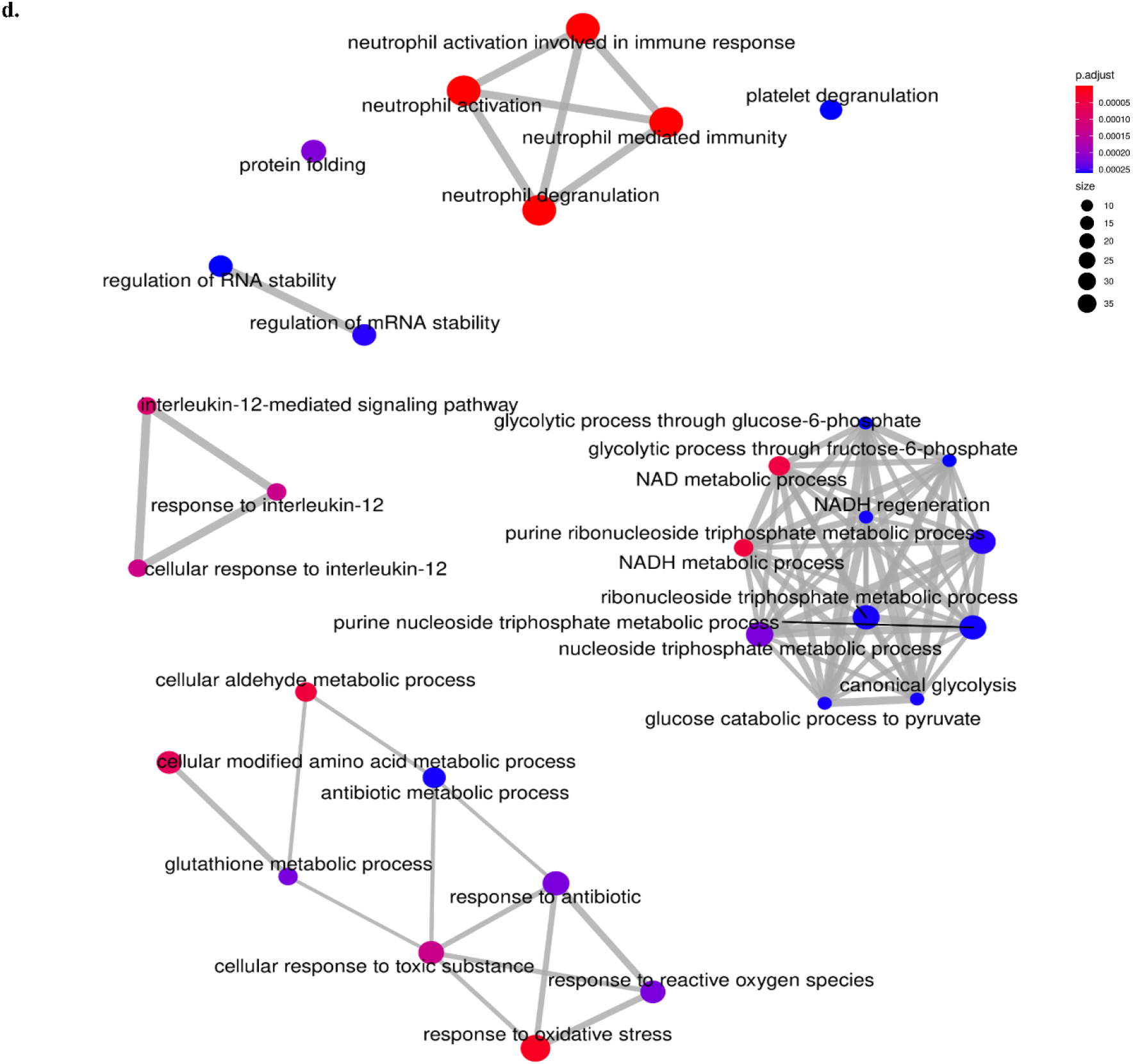
Clinical proteome and characterization of protein dynamics of SARS-COV-2 infected cell. Proteomes of positive samples (clinical swabs). a) Venn diagram of COVID-19 positive and negative host proteome. b) Depicts pathway analysis of unique positive proteome. Dot plot of top 30 pathways according to statistically enriched GO terms are plotted. Y-axis of the plot represents pathways arranged in high to low order of gene counts. c) Category net plot depicting linkages of genes and biological processes as a network for top 4 enriched pathways showing genes involved in them. d) Enrichment map illustrates cluster of functional modules by connecting overlapping gene sets of enriched terms into a network. Emap here, represents overall network and pathways of unique proteome of COVID-19 positive clinical samples. Mainly, GO terms are organised in 5 networks and functional nodules namely with genes involved in protein folding and platelet degranulation are predicted as an individual cluster.

We classified unique proteins from the proteome of positive samples to their GO terms and pathways to characterize host protein dynamics upon viral infection. The statistical significance of GO was further analyzed using the R package, clusterProfiler. In total, proteins unique to positive samples were classified into 244 GO terms. Figure 3B showing a dot plot for the top 30 GO terms according to their statistical significance. We found neutrophil mediated immune responses including degranulation and activation of neutrophil to be higher in positive samples (35 gene count). The abundance of proteins involved in these pathways found to be enriched in positive samples. Additionally, we observed a large collection of proteins involved in the cellular response to oxidative stress and toxic substance and in metabolic pathways, for instance-nucleoside/ribonucleoside triphosphate metabolic process, NAD and NADH metabolic process, amino acid metabolism and glycolytic process. Apart from these, we also observed increased number of proteins involved in RNA processing mechanism including-regulation of RNA/mRNA stability, splicing and localization to Cajal bodies. Among host immune responses, proteins involved in interleukin 12 and 7 mediated signaling pathways were also identified in the positive samples. Figure 3D showing enriched map plot for the pathways identified for unique proteins present in positive samples. Our results indicate that viral infection modulates host in all aspects-biologically, molecularly and cellularly for its survival.

## DISCUSSION

To strengthen the understanding of SARS-CoV-2 in terms of its origin, virulence and host-virus pathogenesis we have analysed the virus genome, proteome as well as host protein response in COVID-19 patients. Our genomic analysis based on these three samples from Bengaluru, which is an urban city in the Southern part of India confirms a high rate of mutation in Indian isolates. Earlier studies observed a low mutation rate of the virus, with an average of 7.23 mutations per sample.^17^ This rate seems to vary between countries whilst India (8.40), Kazakhstan (9.47) and Bangladesh (9.47) show high mutation rate per sample as compared to the world’s average. Whilst India’s average mutation rate is estimated around 8.40, our all three isolates showed ≥11 mutations per sample, with an average mutation rate of 15.33, which is significantly higher than those reported previously. As previous studies were conducted during early phase of pandemic hence, low mutation rate was observed. In total, we found 27 SNPs out of which 14 results in synonymous mutation, 11 missense mutation and 2 are extragenic in nature, observed in the 5’UTR region. 3 of these 11 missense mutations are charged to uncharged or vice-versa amino acid substitution observed in N, S and Orf3a protein. Comparison of the predicted Centroid secondary structure of wild type (WT) and variants (mutant) 5’UTR showing positional entropy of each base, showed there is no change in entropy for 241C>T transition. But entropy is significantly changed for 110C>T transition, not only for that particular base but also for the stem it belongs as compared to WT. This transition may impact viral RNA processing and expression through altered binding of predicted host RBPs-BRUNOL4, RBM38 and SRSF2.^19^ So far, the virus has maintained its genome integrity by avoiding large scale indels and most of the mutations observed are in form of SNPs (Figure 1). The silent mutations could cause a cumulative effect in the long term, in terms of translational efficiency by changing codon usage. Missense mutations may or may not have a direct effect on protein structure and function, but the emergence of new mutations with time may modulate its virulence. Also, mutations in the extragenic region may impact RNA folding, transcription and replication ability.^20^ Based on the marker mutations within the phylogenetic cluster, GISAID introduced a nomenclature system for major clades. Distinct clade wise assignment reveals that all the isolates belong to G derived clade (European origin), isolate 1 to GR and isolate 2 and 3 to GH specifically, highlighting the current prevalence of G originated clades. Though there are 6 major clades currently, more clades can be classified in future with emerging and settling mutations.

To map the evolution and spread of SARS-CoV-2, a global whole-genome phylogenetic tree was created. We observed no direct correlation between isolates and geographic regions though some isolates showed close relations with other isolates of the same country or neighboring countries. For instance, isolates from India mapping close to neighboring countries like Bangladesh, Nepal and China (Hong Kong), stating exchange of virus between neighboring countries. Few of China isolates were found close to Wuhan reference sequence in phylogenetic tree but most of the isolates showed a mosaic pattern of distribution (Figure 2).

Our HRMS data revealed important facets of COVID-19 disease biology. Previous studies also explored this area using viral cell lines, nasopharyngeal swabs and also gargle solution.^21-23^ So far from the clinical swabs, only a few proteins have been identified. Reported studies mainly identified structural proteins of SARS-CoV-2. In this study, we analyzed 12 RT-PCR positive nasopharyngeal swabs for the presence of COVID-19 peptides. In our sample size we found 41 peptides matching to 13 different SARS-CoV-2 proteins. Most of the peptides matched to Nucleocapsid protein and NSP3. 29 peptides matched to ORF1ab polyprotein, 2 to protein 9b, 1 to protein 3a and 9 to structural proteins S (2) and N (11). Detection of ORF9b with two peptides in sample 11 further confirms its expression in clinical samples. Though previous studies predicted function of ORF9b in suppressing Type I and III interferons, ^24,25^ no evidence of its expression was reported so far. This is the first time we are identifying ORF9b in clinical samples which can further be explored to explain the severity of clinical cases and varying susceptibility of individuals. We observed that number of peptides identified did not always correlate with Ct values in the RT-PCR test. ORF10 protein still remains elusive though data showed its role in ubiquitination; its expression in clinical swabs is still a question mark.

In comparison to RNA, proteins are more stable and better candidates for diagnosis. Proteins can be better reflective of virus load and disease. A greater number of peptides identified in sample may indicate greater viral load and proliferation, with samples showing maximum peptides could reflect severity of the disease. Through global proteomics, we identified temporal changes in host proteome upon viral infection. In our sample size, we identified 441 proteins exclusively in positive samples. Pathway analysis of these proteins reveals alteration in basic host processes and heightened immune response (Figure 4), as also reported previously, which showed that host cells try to combat viral load by alleviating immune response especially, mediated by macrophage, complement and IL-6 signaling.^26,27^ Here we observed enrichment of neutrophil-mediated immune responses which are known to play a crucial role in airway infections and elicit antiviral response.^28^ Not many studies are there who commented on the role of neutrophils in COVID-19 and more or less their role is not very clear. Though they are important for effective immune response, they can also be cytotoxic and lead to hyperinflammation through degranulation and lysis during severe pneumonia.^28-30^ With increasing studies showing upregulated neutrophil genes and neutrophil-attracting chemokines in SARS-CoV-2, current literature also showed their role in damaging host inflammatory responses through involvement of NETs and increased neutrophil-to-lymphocyte ratio observed in severe cases, indicating their association to COVID-19 pathology.^31-33^ Hence, further studies would be useful in dissecting their role and to understand the mechanism involved in severe cases.

We also observed more proteins involved in the cellular response to IL-12 and IL-7, which has a role in adaptive immune system mainly T-cell mediated immune response. As expected, we observed a heightened response to oxidative stress. An additional cluster of proteins identified in clinical swabs mainly enriched in RNA processing (splicing, spliceosomal components), mRNA stability, localization to Cajal body and RNA metabolism. This result relates to previous studies which also suggested splicing as a crucial pathway for SARS-CoV-2 survival.^21,34^ Pathways involved in metabolism, for instance-Carbon metabolism, RNA/DNA synthesis, NAD/NADH synthesis, unsaturated fatty acid metabolism also among the enriched pathways observed in positive samples. Unsaturated fatty acid are components of phospholipids and involves in maintenance of membrane fluidity. Phospholipids along with sphingolipids mediate signal transduction and immune responses. Earlier studies reported phagocytosis and platelet degranulation mediated alteration in production of glycerophospholipid, and reduction of glycerophospholipid upon SARS-CoV-2 infection.^35,26^ Overall, through our study we suggest a complete alteration of host processes affecting cellular, metabolic or biological functions. Furthermore, our global proteomics unravels cellular and molecular pathways for therapeutic interventions. Essentially, our result suggests that proteomics can not only offer timely and sensitive diagnosis but also has the ability to reveal prognosis and severity of viral infection.

Altogether our study combining genomics and proteomics revealed genome-wide SNPs, COVID-19 proteome and dynamics of host proteome in clinical samples. In combination with etiological and patient severity details, multiomics studies cannot only predict the progression of SARS-CoV-2 but will also help in identifying drug targets to offer counter treatment for this as well as for future pandemics.

### CONCLUSION

In this proteo-genomic study of SARS-CoV-2, we analyzed the clinical proteome of COVID-19 and the variations accumulated in the genome since its identification. The clinical landscape of SARS-CoV-2 and host proteome highlighted correlation between the viral proteins and host responses (Figure 5). Through our proteomics study, we confirmed expression of various (13 in this study) viral proteins in the host cell. Pathway analysis of host proteome indicated enrichment of proteins majorly involved in immune response, metabolism and RNA processing. We identified several COVID-19 peptides within a 90 min MS-acquisition window, few of which are unique to SARS-CoV-2 and not present in other coronaviruses, confirming SARS-CoV-2 infection. Our study suggests the importance of SARS-CoV-2 specific peptide LVDPQIQLAVTR (Orf9b) and IFTIGTVTLKQGEIK (Orf3a) in its accurate detection. Though more studies can further deepen the understanding of viral biopathology, this study offers both proteo-genomic analysis of SARS-CoV-2 confirming high rate of mutation in Indian isolates and expression of viral protein (Orf9b) in the host cell, which suppresses the host innate immune response. Enrichment of proteins involved in neutrophils mediated immune response pointed towards the crosstalk between host and pathogen. Our study highlighted the potential of mass-spectrometry as a specific and sensitive diagnostic tool and laid down the foundation for future studies. Further studies combined with patient severity details can help in predicting the prognosis of viral infection.

**Figure 5.**
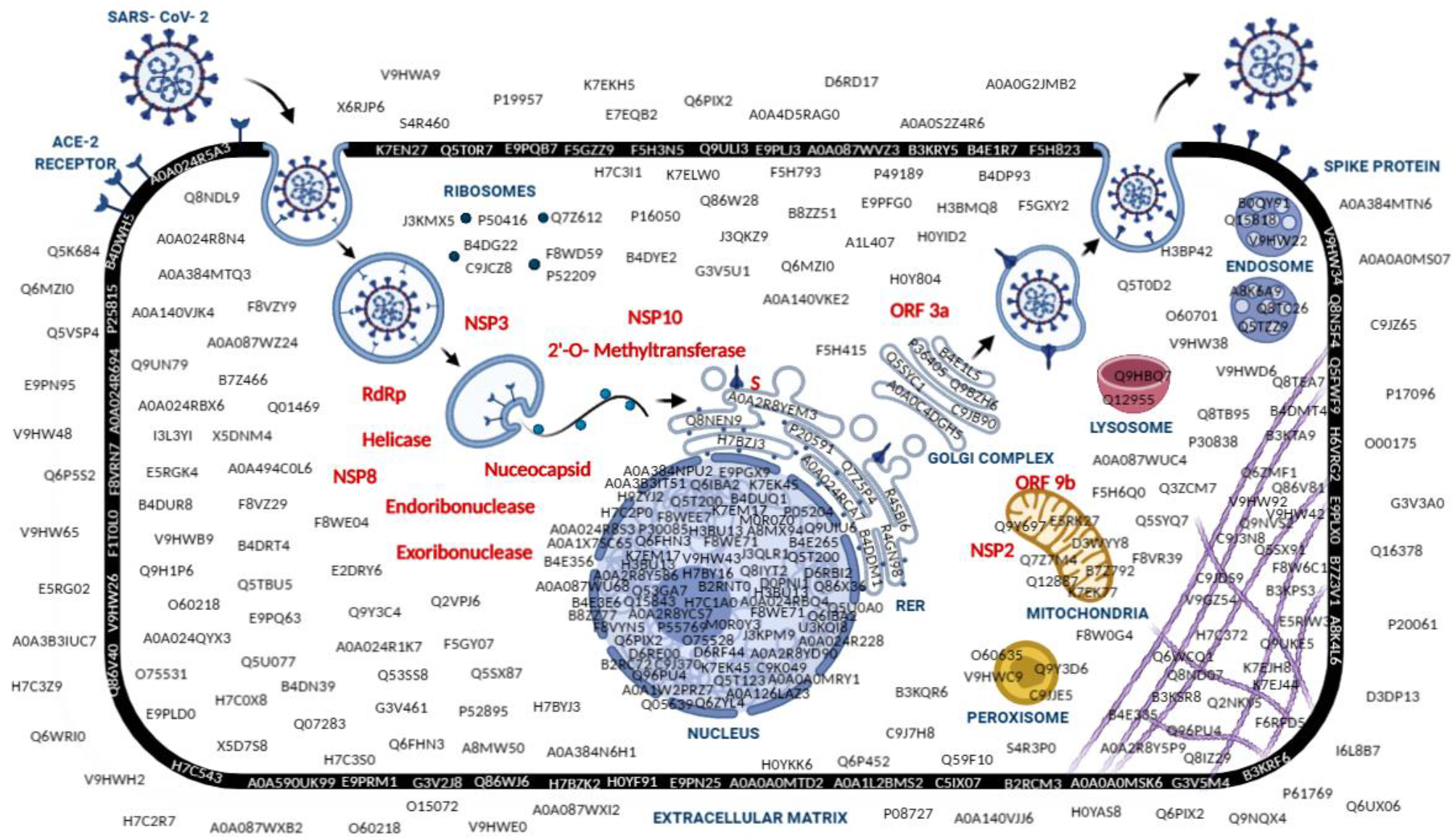
Schematic representation of clinical proteome of COVID-19 patient. The figure depicts COVID-19 and host-epithelial cell proteomes identified in this study. Upon vesicular internalization, the coronavirus RNA undergoes a series of replication and translates into viral proteins. Viral proteins are indicated in red text while host proteins identified in positive samples are shown in black. Viral structural proteins like spike glycoprotein undergoes folding and trafficking through ER. The nucleocapsid form (genome and N) individually assembled in the cytoplasm further joins structural proteins in ER-Golgi intermediate compartment forming a complete virion particle. The assembled virion particles exit the infected cell by exocytosis and continue to transmit.

## Data Availability

Mass spectrometry proteomics data acquired on nasopharyngeal swabs have been deposited
to the ProteomeXchange Consortium via the PRIDE15 partner repository under dataset identifiers PXD021896 and 10.6019/PXD021896.
The SARS-CoV-2 whole genome sequences have been submitted to NCBI under the accession number PRJNA668889.

## AUTHOR INFORMATION

### Corresponding Author

*Utpal Tatu-Department of Biochemistry, Indian Institute of Science, Bangalore - 560012, India., Telephone: +91(080) 22932823; Fax: 23600814

### Authors

Sheetal Tushir-Department of Biochemistry, Indian Institute of Science, Bangalore - 560012 Sathisha Kamanna-Department of Biochemistry, Indian Institute of Science, Bangalore - 560012 Sujith S Nath-Department of Biochemistry, Indian Institute of Science, Bangalore - 560012 Aishwarya Bhat-Department of Biochemistry, Indian Institute of Science, Bangalore - 560012 Steffimol Rose-Department of Biochemistry, Indian Institute of Science, Bangalore- 560012 Advait R Aithal-Department of Biochemistry, Indian Institute of Science, Bangalore- 560012

## Author Contributions

‡ST and SK contributed equally. ST analyzed genomic and proteomic data. SK performed the mass-spectrometry work. SK and ST analyzed the mass-spectrometry data. SN and ST performed mass-spectrometry sample preparation. AB contributed to phylogenetic and genomic analysis. SR and ST performed pathway analysis for proteomic data. AA coordinated the clinical COVID-19 samples. UT conceived the idea of this study. UT and ST wrote the manuscript.

### Notes

The authors declare no conflict of interest.

The mass spectrometry and proteomics data acquired on clinical nasopharyngeal swabs have been deposited to the ProteomeXchange Consortium via the PRIDE partner repository with the dataset identifiers PXD021896 and 10.6019/PXD021896.

## ACKNOWLEDGMENTS

Authors thank funding for the study by Department of Biotechnology and DBT-IISc Partnership grant. ST acknowledge fellowship from IISc. SK acknowledge fellowship from DBT-IISc Postdoctoral Research Associateship. Authors acknowledge Agilent technologies for their collaboration based on LC/Q-TOF and technical assistance. Dr. Anup Chugani is acknowledged for his assistance in genome sequencing and analysis.

## ABBREVIATIONS

SARS-CoV-2: Severe acute respiratory syndrome coronavirus 2
COVID-19: Coronavirus disease 2019
RT-PCR: Reverse transcription-polymerase chain reaction
GISAID: Global Initiative on Sharing All Influenza Data
HRMS: High-resolution mass spectrometry
NGS: Next-generation sequencing
SNPs: Single Nucleotide Polymorphism
GO: Gene Ontology

---

**For TOC Only**

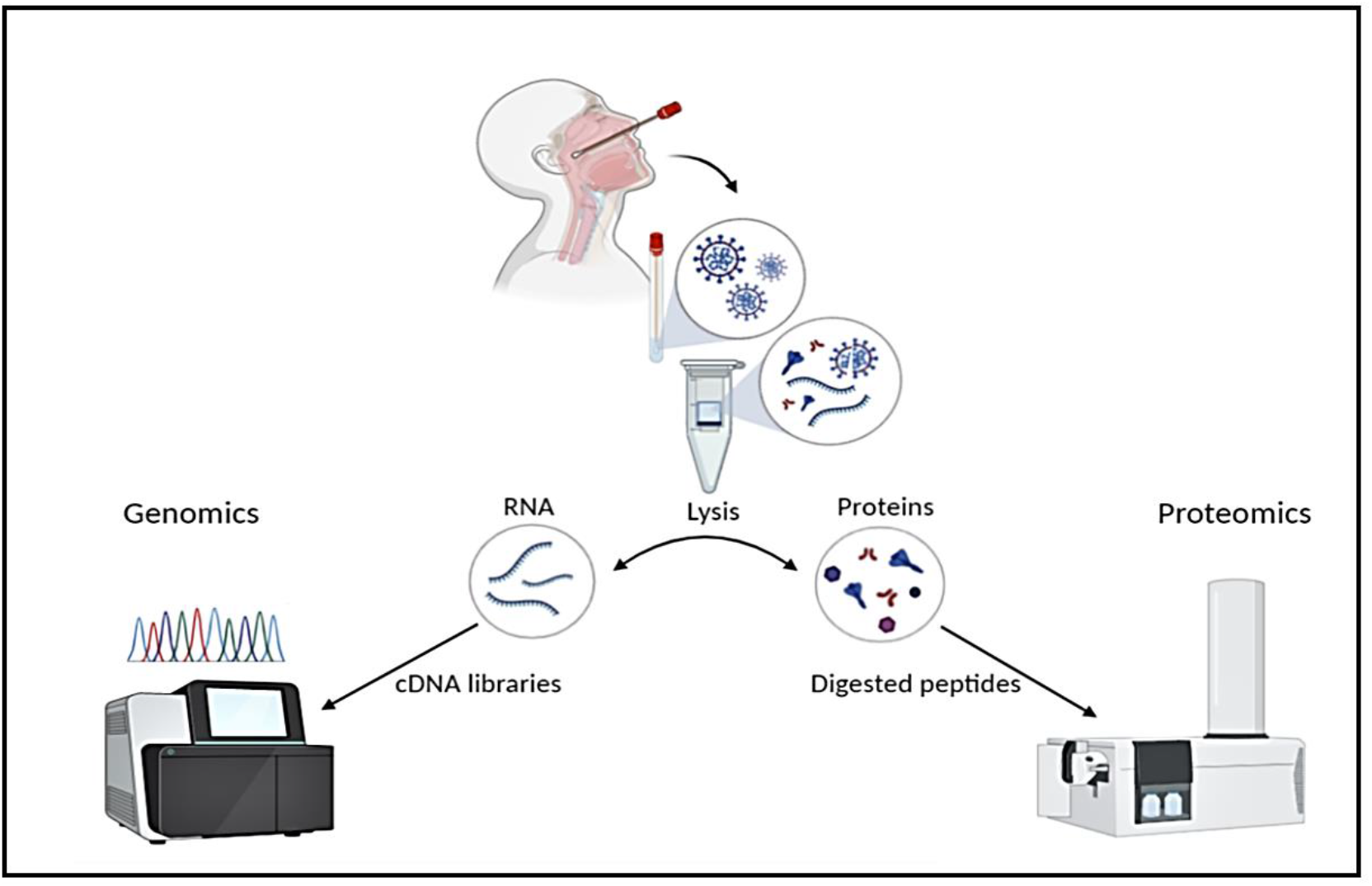

## REFERENCES

1. Drosten, C., Günther, S., Preiser, W., van der Werf, S., Brodt, H. R., Becker, S., Rabenau, H., Panning, M., Kolesnikova, L., Fouchier, R. A., Berger, A., Burguière, A. M., Cinatl, J., Eickmann, M., Escriou, N., Grywna, K., Kramme, S., Manuguerra, J. C., Müller, S., Rickerts, V., Stürmer, M., Vieth, S., Klenk, H. D., Osterhaus, A. D., Schmitz, H., Doerr, H. W., Identification of a novel coronavirus in patients with severe acute respiratory syndrome. N. Engl. J. Med. 2003, 348 (20), 1967–76.

2. Ksiazek, T. G., Erdman, D., Goldsmith, C. S., Zaki, S. R., Peret, T., Emery, S., Tong, S., Urbani, C., Comer, J. A., Lim, W., Rollin, P. E., Dowell, S. F., Ling, A. E., Humphrey, C. D., Shieh, W. J., Guarner, J., Paddock, C. D., Rota, P., Fields, B., DeRisi, J., Yang, J. Y., Cox, N., Hughes, J. M., LeDuc, J. W., Bellini, W. J., Anderson, L. J., A novel coronavirus associated with severe acute respiratory syndrome. N. Engl. J. Med. 2003, 348 (20), 1953–66.

3. Zaki, A. M., van Boheemen, S., Bestebroer, T. M., Osterhaus, A. D., Fouchier, R. A., Isolation of a novel coronavirus from a man with pneumonia in Saudi Arabia. N. Engl. J. Med. 2012, 367 (19), 1814–20.

4. (WHO), W. H. O. Report of the WHO-China Joint Mission on Coronavirus Disease 2019 (COVID-19) https://www.who.int/docs/default-source/coronaviruse/who-china-joint-mission-on-covid-19-final-report.pdf.

5. Wu, F., Zhao, S., Yu, B., Chen, Y. M., Wang, W., Song, Z. G., Hu, Y., Tao, Z. W., Tian, J. H., Pei, Y. Y., Yuan, M. L., Zhang, Y. L., Dai, F. H., Liu, Y., Wang, Q. M., Zheng, J. J., Xu, L., Holmes, E. C., Zhang, Y. Z., A new coronavirus associated with human respiratory disease in China. Nature 2020, 579 (7798), 265–269.

6. Durrant, J. D., Kochanek, S. E., Casalino, L., Ieong, P. U., Dommer, A. C., Amaro, R. E., Mesoscale All-Atom Influenza Virus Simulations Suggest New Substrate Binding Mechanism. ACS Cent. Sci. 2020, 6 (2), 189–196.

7. Leggett, R., Ramirez-Gonzalez, R., Clavijo, B., Waite, D., Davey, R., Sequencing quality assessment tools to enable data-driven informatics for high throughput genomics. Front. Genet. 2013, 4 (288).

8. Martin, M., Cutadapt removes adapter sequences from high-throughput sequencing reads. 2011 2011, 17 (1), 3.

9. Severe acute respiratory syndrome coronavirus 2 isolate Wuhan-Hu-1, complete genome. https://www.ncbi.nlm.nih.gov/nuccore/NC_045512.2?report=genbank.

10. Li, H., Durbin, R., Fast and accurate long-read alignment with Burrows–Wheeler transform. Bioinformatics 2010, 26 (5), 589–595.

11. Brouard, J.-S., Schenkel, F., Marete, A., Bissonnette, N., The GATK joint genotyping workflow is appropriate for calling variants in RNA-seq experiments. J. Anim. Sci. Biotechnol. 2019, 10 (1), 44.

12. Cingolani, P., Platts, A., Wang le, L., Coon, M., Nguyen, T., Wang, L., Land, S. J., Lu, X., Ruden, D. M., A program for annotating and predicting the effects of single nucleotide polymorphisms, SnpEff: SNPs in the genome of Drosophila melanogaster strain w1118; iso-2; iso-3. Fly (Austin) 2012, 6 (2), 80–92.

13. Tamura, K., Nei, M., Estimation of the number of nucleotide substitutions in the control region of mitochondrial DNA in humans and chimpanzees. Mol. Biol. Evol. 1993, 10 (3), 512–26.

14. Kumar, S., Stecher, G., Li, M., Knyaz, C., Tamura, K., MEGA X: Molecular Evolutionary Genetics Analysis across Computing Platforms. Mol. Biol. Evol. 2018, 35 (6), 1547–1549.

15. Perez-Riverol, Y., Csordas, A., Bai, J., Bernal-Llinares, M., Hewapathirana, S., Kundu, D. J., Inuganti, A., Griss, J., Mayer, G., Eisenacher, M., Pérez, E., Uszkoreit, J., Pfeuffer, J., Sachsenberg, T., Yilmaz, S., Tiwary, S., Cox, J., Audain, E., Walzer, M., Jarnuczak, A. F., Ternent, T., Brazma, A., Vizcaíno, J. A., The PRIDE database and related tools and resources in 2019: improving support for quantification data. Nucleic Acids Res. 2019, 47 (D1), D442–d450.

16. GISAID. https://www.gisaid.org/.

17. Mercatelli, D., Giorgi, F. M., Geographic and Genomic Distribution of SARS-CoV-2 Mutations. Front. Microbiol. 2020, 11 (1800).

18. Proteomes. https://www.uniprot.org/proteomes/?query=sars+cov-2&sort=score.

19. Mukherjee, M., Goswami, S., Global cataloguing of variations in untranslated regions of viral genome and prediction of key host RNA binding protein-microRNA interactions modulating genome stability in SARS-CoV-2. PloS one 2020, 15 (8), e0237559–e0237559.

20. Kim, D., Lee, J.-Y., Yang, J.-S., Kim, J. W., Kim, V. N., Chang, H., The Architecture of SARS-CoV-2 Transcriptome. Cell 2020, 181 (4), 914-921.e10.

21. Grenga, L., Gallais, F., Pible, O., Gaillard, J.-C., Gouveia, D., Batina, H., Bazaline, N., Ruat, S., Culotta, K., Miotello, G., Debroas, S., Roncato, M.-A., Steinmetz, G., Foissard, C., Desplan, A., Alpha-Bazin, B., Almunia, C., Gas, F., Bellanger, L., Armengaud, J., Shotgun proteomics analysis of SARS-CoV-2-infected cells and how it can optimize whole viral particle antigen production for vaccines. Emerg. Microbes Infect. 2020, 9 (1), 1712–1721.

22. Gouveia, D., Miotello, G., Gallais, F., Gaillard, J.-C., Debroas, S., Bellanger, L., Lavigne, J.-P., Sotto, A., Grenga, L., Pible, O., Armengaud, J., Proteotyping SARS-CoV-2 Virus from Nasopharyngeal Swabs: A Proof-of-Concept Focused on a 3 Min Mass Spectrometry Window. J. Proteome Res. 2020.

23. Ihling, C., Tänzler, D., Hagemann, S., Kehlen, A., Hüttelmaier, S., Arlt, C., Sinz, A., Mass Spectrometric Identification of SARS-CoV-2 Proteins from Gargle Solution Samples of COVID-19 Patients. J. Proteome Res. 2020.

24. Jiang, H.-w., Zhang, H.-n., Meng, Q.-f., Xie, J., Li, Y., Chen, H., Zheng, Y.-x., Wang, X.-n., Qi, H., Zhang, J., Wang, P.-H., Han, Z.-G., Tao, S.-c., SARS-CoV-2 Orf9b suppresses type I interferon responses by targeting TOM70. Cell. Mol. Immunol. 2020, 17 (9), 998–1000.

25. Han, L., Zhuang, M.-W., Zheng, Y., Zhang, J., Nan, M.-L., Wang, P.-H., Gao, C., SARS-CoV-2 ORF9b Antagonizes Type I and III Interferons by Targeting Multiple Components of RIG-I/MDA-5-MAVS, TLR3-TRIF, and cGAS-STING Signaling Pathways. bioRxiv 2020, 2020.08.16.252973.

26. Shen, B., Yi, X., Sun, Y., Bi, X., Du, J., Zhang, C., Quan, S., Zhang, F., Sun, R., Qian, L., Ge, W., Liu, W., Liang, S., Chen, H., Zhang, Y., Li, J., Xu, J., He, Z., Chen, B., Wang, J., Yan, H., Zheng, Y., Wang, D., Zhu, J., Kong, Z., Kang, Z., Liang, X., Ding, X., Ruan, G., Xiang, N., Cai, X., Gao, H., Li, L., Li, S., Xiao, Q., Lu, T., Zhu, Y., Liu, H., Chen, H., Guo, T., Proteomic and Metabolomic Characterization of COVID-19 Patient Sera. Cell 2020, 182 (1), 59-72.e15.

27. Messner, C. B., Demichev, V., Wendisch, D., Michalick, L., White, M., Freiwald, A., Textoris-Taube, K., Vernardis, S. I., Egger, A.-S., Kreidl, M., Ludwig, D., Kilian, C., Agostini, F., Zelezniak, A., Thibeault, C., Pfeiffer, M., Hippenstiel, S., Hocke, A., von Kalle, C., Campbell, A., Hayward, C., Porteous, D. J., Marioni, R. E., Langenberg, C., Lilley, K. S., Kuebler, W. M., Mülleder, M., Drosten, C., Suttorp, N., Witzenrath, M., Kurth, F., Sander, L. E., Ralser, M., Ultra-High-Throughput Clinical Proteomics Reveals Classifiers of COVID-19 Infection. Cell Systems 2020, 11 (1), 11-24.e4.

28. Camp, J. V., Jonsson, C. B., A Role for Neutrophils in Viral Respiratory Disease. Front. in Immunol. 2017, 8 (550).

29. Haick, A. K., Rzepka, J. P., Brandon, E., Balemba, O. B., Miura, T. A., Neutrophils are needed for an effective immune response against pulmonary rat coronavirus infection, but also contribute to pathology. J. General Virol. 2014, 95 (3), 578–590.

30. Zhu, B., Zhang, R., Li, C., Jiang, L., Xiang, M., Ye, Z., Kita, H., Melnick, A. M., Dent, A. L., Sun, J., BCL6 modulates tissue neutrophil survival and exacerbates pulmonary inflammation following influenza virus infection. Proc. Natl. Acad. Sci. 2019, 116 (24), 11888–11893.

31. Veras, F. P., Pontelli, M., Silva, C., Toller-Kawahisa, J., de Lima, M., Nascimento, D., Schneider, A., Caetite, D., Rosales, R., Colon, D., Martins, R., Castro, I., Almeida, G., Lopes, M. I., Benatti, M., Bonjorno, L., Giannini, M., Luppino-Assad, R., Almeida, S., Vilar, F., Santana, R., Bollela, V., Martins, M., Miranda, C., Borges, M., Pazin-Filho, A., Cunha, L., Zamboni, D., Dal-Pizzol, F., Leiria, L., Siyuan, L., Batah, S., Fabro, A., Mauad, T., Dolhnikoff, M., Duarte-Neto, A., Saldiva, P., Cunha, T., Alves-Filho, J. C., Arruda, E., Louzada-Junior, P., Oliveira, R., Cunha, F., SARS-CoV-2 triggered neutrophil extracellular traps (NETs) mediate COVID-19 pathology. medRxiv 2020, 2020.06.08.20125823.

32. Barnes, B. J., Adrover, J. M., Baxter-Stoltzfus, A., Borczuk, A., Cools-Lartigue, J., Crawford, J. M., Daßler-Plenker, J., Guerci, P., Huynh, C., Knight, J. S., Loda, M., Looney, M. R., McAllister, F., Rayes, R., Renaud, S., Rousseau, S., Salvatore, S., Schwartz, R. E., Spicer, J. D., Yost, C. C., Weber, A., Zuo, Y., Egeblad, M., Targeting potential drivers of COVID-19: Neutrophil extracellular traps. J. Exp. Med. 2020, 217 (6).

33. Liu, J., Liu, Y., Xiang, P., Pu, L., Xiong, H., Li, C., Zhang, M., Tan, J., Xu, Y., Song, R., Song, M., Wang, L., Zhang, W., Han, B., Yang, L., Wang, X., Zhou, G., Zhang, T., Li, B., Wang, Y., Chen, Z., Wang, X., Neutrophil-to-lymphocyte ratio predicts critical illness patients with 2019 coronavirus disease in the early stage. J. Transl. Med. 2020, 18 (1), 206.

34. Bojkova, D., Klann, K., Koch, B., Widera, M., Krause, D., Ciesek, S., Cinatl, J., Münch, C., Proteomics of SARS-CoV-2-infected host cells reveals therapy targets. Nature 2020, 583 (7816), 469–472.

35. Rouzer, C. A., Ivanova, P. T., Byrne, M. O., Brown, H. A., Marnett, L. J., Lipid profiling reveals glycerophospholipid remodeling in zymosan-stimulated macrophages. Biochemistry 2007, 46 (20), 6026–42.

